# Association between body roundness index and myopia among US adolescents: a cross-sectional study of NHANES 2001-2006

**DOI:** 10.1101/2025.01.05.25320015

**Authors:** Dongfang Wang, Fang Sha, JiaoJiao Feng, Weihua Su, Guodong Tang, Jike Song, Hongsheng Bi

## Abstract

**Background:** This study analyzed the association between body roundness index (BRI) and myopia in adolescents in the United States.

**Methods:** This cross-sectional study analyzed the data of a nationally representative survey, the National Health and Nutrition Examination Survey (NHANES) conducted from 2001 to 2006. Among the 7078 adolescents aged 12 to 19 years, 3915 participants were selected for review. BRI, a new obesity assessment index that evaluates visceral fat, was classified into four groups: Q1,< 2.083; Q2, 2.083 to 2.724; Q3, 2.725 to 4.004; Q4, ≥ 4.005. Myopia was defined as SE≤-0.50 diopters (D). Weighted multivariate logistic regression analysis and smooth curve fitting were performed to evaluate the association between BRI and myopia. Additionally, subgroup and sensitivity analysis was applied.

**Results:** The prevalence of myopia was 39.4%. Adolescents who are older, have a larger waist circumference, a lower level of vitamin D, a relatively low PIR level, and a higher waist-height ratio (WHtR) are more likely to have a higher BRI. We found a positive association between BRI and myopia before and after adjusting for confounders (odds ratio [OR] = 1.057, 95% confidence interval [CI]: 1.016-1.100, *P* < 0.01; OR = 1.049, 95%CI: 1.009-1.090, *P*=0.022). Specifically, after full adjustment for age, sex, race/ethnicity, PIR, guardians’ education, Vitamin D, physical activity, screen time, adolescents in Q4 were 26.1% more likely to develop myopia compared to adolescents in Q1 (OR=1.261, 95% CI:1.046-1.521, *P*=0.022). There was a general linear trend between BRI and myopia (all *P* values for trend <0.001) and nonlinear association (all *P* for nonlinear < 0.05). Subgroup analysis conducted after full adjustment demonstrated positive associations between BRI and myopia in Mexican American adolescents (OR=1.11, 95% CI: 1.05-1.19, *P*=0.001), women (OR=1.10, 95% CI: 1.05-1.15, *P*<0.001), those guardians with educational levels lower than high school (OR=1.09, 95% CI: 1.03-1.14, *P*=0.004), those with PIR < 1.30 (OR=1.07, 95% CI:1.01-1.14, *P*=0.037) and those with high physical activity (OR=1.08, 95% CI: 1.04-1.13, *P*<0.001). Sensitivity analysis was applied using weighted ordinal logistic regressions to explore the relationship of BRI with degrees of myopia. The results remained stable after adjusting for potential confounding factors, consistent with the results of BRI and myopia.

**Conclusions:** This study assessed that in adolescents, an increase in BRI was associated with an increased risk of myopia, especially in women. Additionally, there was a nonlinear association between BRI and myopia. This study aimed to increase public awareness of BRI values, a novel measure of obesity, and that maintaining a moderate BRI can help reduce the risk of myopia.

## 1. Introduction

Myopia has become a major public health problem worldwide. The number of myopias has been increasing in recent decades. It is estimated that myopia will affect half of the world’s population (about 4758 million people) by 2050 [1]. In China, the overall prevalence of myopia among children and adolescents is 52.7%, with a rate of 71.1% among junior high school students and 80.5% among senior high school students [2]. Myopia, particularly high myopia, can increase the risk of ocular pathologies such as retinal detachment, cataract, glaucoma, staphyloma, myopic macular degeneration, and myopic choroidal neovascularization [3, 4], resulting in irreversible vision loss and serious influence on daily life. Therefore, it is essential to prevent myopic individuals from developing high myopia.

Obesity is a global epidemic with high prevalence and contributes to many health issues [5]. The prevalence of obesity in Chinese students aged 7–18 years increased from 0.13% in 1985 to 7.26% in 2014 [6]. There are many indicators representing obesity, such as body mass index (BMI), waist circumference (WC), and waist-height ratio (WHtR), among which BMI is the most representative measure. However, these traditional measures of obesity have limitations. For example, BMI, representing overall obesity, does not measure the specific distribution of body fat, and WC and WHtR do not distinguish between visceral and subcutaneous fat. As body composition research has been extensively conducted, more attention has been paid to visceral obesity[7, 8]. Thomas et al. proposed the Body Roundness Index (BRI), an innovative obesity-related human body measurement indicator, to provide a relatively more accurate measurement method to assess body fat and visceral obesity [9]. The BRI, affected by height and waist circumference, can more comprehensively reflect the distribution of visceral fat. BRI has been shown to be more effective than other anthropometric measures in assessing the risk associated with a variety of clinical outcomes, including cardiometabolic disease[10–12], cancer [13], and all-cause mortality [14]. Therefore, we chose to study the relationship between BRI and myopia instead of that between BMI and myopia.

The association between obesity and myopia has been studied in recent decades [15, 16]. In previous studies, obesity was measured by body mass index (BMI) and several studies have demonstrated that obesity defined by BMI was associated with myopia. A study of NHANES reported a positive linear relationship between BMI and myopia (OR 1.01; 95% CI 1.01-1.02; p<0.05) [17]. In a nationwide cross-sectional study of more than 1.3 million adolescents, myopia was higher in underweight individuals, lowest in those with healthy weight, and then increased with increasing BMI category, in a pattern J-shaped [18]. Similarly, a Korean study of 938 children aged 5 to 18 years found that a higher BMI was significantly associated with high myopia [19]. However, a British cohort study among 2,487 examinees reported that a lower BMI at the prepubertal and postpubertal ages was associated with more severe and early-onset myopia [20]. To our knowledge, no research has been conducted on the correlation between BRI and myopia. To fill this gap, we aim to explore the relationship between BRI and myopia.

## 2. Methods

### 2.1 Study population

The National Health and Nutrition Examination Survey (NHANES) is an ongoing series of nationally representative surveys initiated by the Centers for Disease Control and Prevention (CDC). Since 1999, NHANES has been conducted in 2-year cycles. Every year, approximately 5,000 people from 15 counties are surveyed by in-home interviews and a mobile examination center (MEC). Its primary objective was to examine the health and nutritional status of children and adults throughout America by collecting data on demographic, socioeconomic and health-related variables, including physiological and laboratory measurements.

This database can be accessed on the official website of NHANES: www.cdc.gov/nchs/NHANES. NHANES information is available and free to the public, and all participants and/or their guardians have provided their written informed consent. The NHANES study protocol has been approved by the Ethics Review Board of the National Center for Health Statistics.

We extracted information from 7078 adolescents from 3 independent cycles (2001-2002,2003-2004,2005-2006) of the NHANES. Of these participants, we further excluded 3163 individuals due to missing relevant information. The final sample size was 3915 adolescents. The detailed inclusion and exclusion processes are presented in Fig 1.

**Figure 1.**
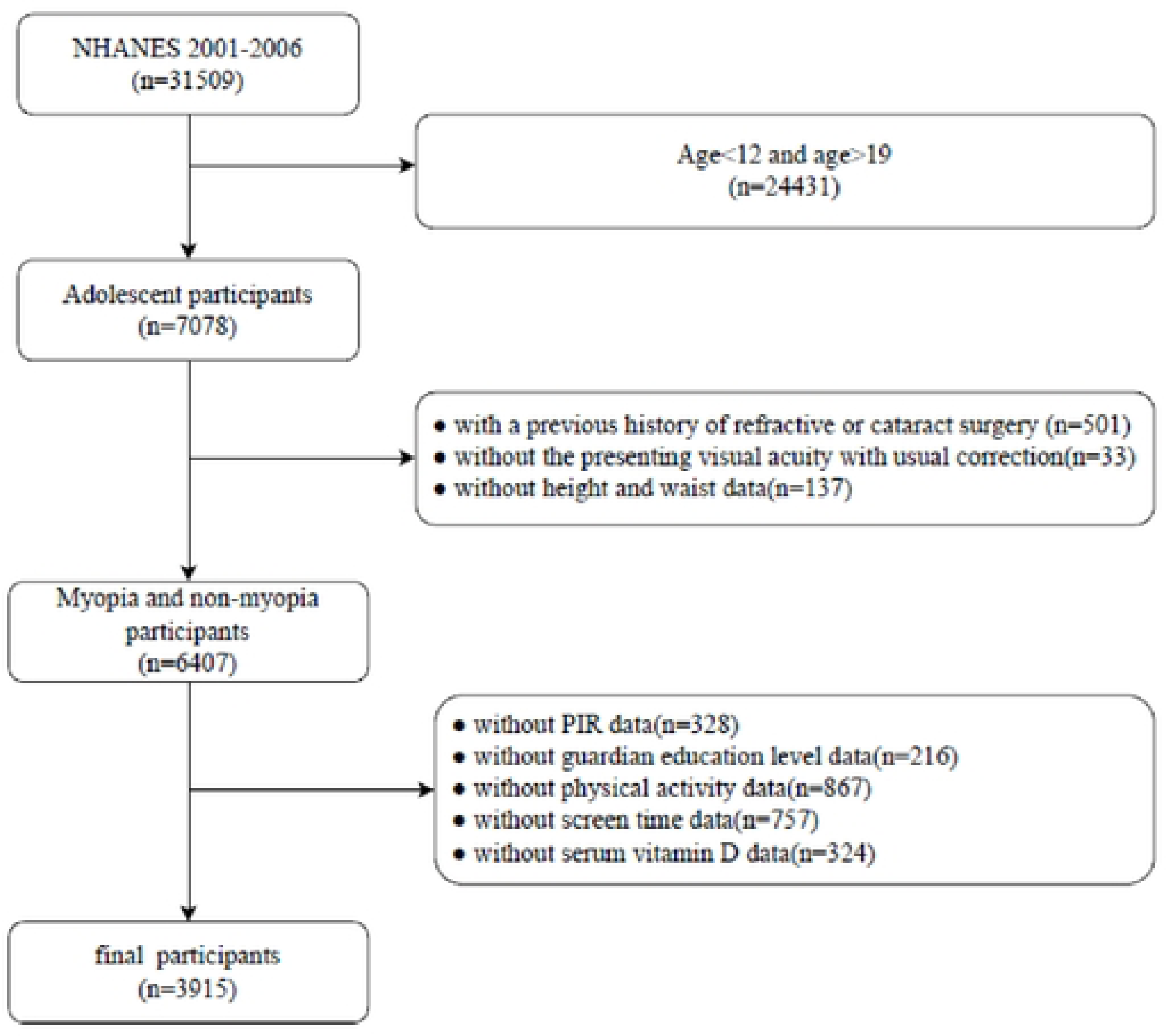
Flow chart of the sample selection process.

The inclusion criteria were adolescents between the ages of 12 and 19 years old.

The exclusion criteria were as follows:

1. subjects with a history of refractive or cataract surgery;
2. subjects without or unavailable data on spherical and cylindrical dioptric power;
3. subjects without height and waist data;
4. subjects lacking covariate data.

### 2.2 Evaluation of myopia

In this study, participants underwent a non-cycloplegic vision examination at the Mobile Examination Center (MEC). Due to the high correlation of refractive errors between the right and left eyes, we used only the right eye as the evaluation eye. The objective refraction (sphere and cylinder) results were acquired by the median of three consecutive measurements using an autorefractor (Nidek ARK-760 A, Nidek Co. Ltd., Gamagori, Japan). The spherical equivalent (SE) was calculated by the sphere+1/2 cylinder. Myopia was defined as a SE≤-0.50 diopters (D). Specifically, myopia was categorized into mild (−3.00 D<SE≤-0.50 D), moderate (−6.00D<SE ≤-3.00 D), or high (SE≤-6.00 D) myopia[21, 22](S1 Table).

### 2.3 Evaluation of BRI

Body roundness index (BRI) is a new obesity assessment index evaluating visceral fat that measures body height (cm) and waist circumference (cm) (WC). In the NHANES database, height and WC were measured in MEC by trained health technicians. The BRI is calculated using the following formula [9]:

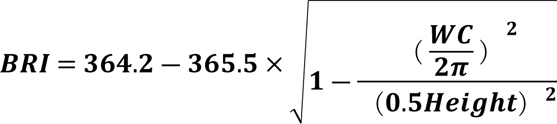

Due to the lack of the reference range, the BRI was grouped by weighted quartiles to explore its correlation with adolescent myopia.

### 2.4 Covariates

In order to control the potential confounder variables, we included the following covariates in this study. Age, sex(male, female), race and ethnicity, PIR(family poverty income ratio, educational levels of guardians were collected through in-home interviews as demographic covariates. Serum vitamin D was extracted from laboratory data. Physical activity and screen time (hours per day) were obtained through the questionnaire data. All detailed measurement procedures of the variables in this study are publicly accessible at http://www.cdc.gov/nchs/nhanes/.

#### 2.4.1 Race

Race was divided by NHANES into five categories: Mexican American, other Hispanic, non-Hispanic white, non-Hispanic black, and other races [23].

#### 2.4.2 Educational levels of guardians

The educational levels of the guardians were classified as less than high school, high school and more than high school.

#### 2.4.3 PIR

The poverty income ratio (PIR) is used to assess income status and is calculated by dividing the total household income by the annual poverty threshold as determined by the U.S. Census Bureau based on household size. A PIR<1 indicates that family income was below the poverty threshold [24]. Three categories were created for household income: low (PIR ≤1.30), middle (PIR>1.30 to ⩽3.50), and high (PIR >3.50) [25].

#### 2.4.4 Physical activity

Physical activity was assessed using a Physical Activity Questionnaire (PAQ) that included questions related to all physical activities performed within the past 30 days, involving activity type, duration, intensity and frequency. Moderate intensity activity was considered to lead to slight sweating or a mild to moderate elevation in breathing or heart rate (HR), while vigorous activity was considered to trigger heavy sweating or a significant increase in breathing or HR. Metabolic equivalent scores were calculated based on activity type and intensity [26, 27]. To obtain the MET minutes per 30 days (MET min/30d) for each activity, the MET score was multiplied by the average duration and frequency of performance in the past 30 days. Subsequently, the MET min/30d of each activity was summed and then divided by 4.29 to determine the total MET minutes per week. Participants were categorized into low and high physical activity groups according to whether they adhered to the national physical activity guidelines, with low physical activity less than 500 MET/week and high physical activity greater than or equal to 500 MET/week [28].

#### 2.4.5 Screen time

Television/video viewing was evaluated by the question: ‘‘Over the past 30 days, on average about how many hours per day did you sit and watch TV? Would you say… less than 1 hour, 1 hour, 2 hours, 3 hours, 4hours, 5 hours or more, or none?” Computer use was assessed similarly using the question: ‘‘Over the past 30 days, on average about how many hours per day did you use a computer or play computer games? Would you say… less than 1 hour, 1 hour, 2 hours,3 hours, 4 hours, 5 hours or more, or none?’’ The average number of hours the participants spent sitting and watching TV and the average number of hours using a computer or playing computer games were combined, and a dichotomous variable was created based on the recommendations of the American Academy of Pediatrics (AAP) (a maximum of 2 hours of screen time per day). Participants who met the recommendation were classified as having low screen time and those who did not were classified as having high screen time [29].

### 2.5 Statistical analyses

Taking into account the complex sampling design of the NHANES, statistical analyses were performed following the CDC guidelines and using appropriate weights for the sampling of the NHANES [30].

Weighted continuous measurement data in accordance with the normal distribution were described by mean (SE), while median(Q1, Q3) were used to describe data that did not comply with the normal distribution. Categorical data were described by n (%). Weighted analysis of variance and Pearson’s chi-square tests were used to evaluate the differences in baseline characteristics among the BRI quartiles. Weighted univariate and multivariate logistic regression analysis was performed to evaluate the association between BRI and myopia. Model 1 was the unadjusted model. Model 2 was adjusted for age, sex, race and ethnicity. Model 3 was fully adjusted for age, sex, race and ethnicity, the educational level of guardians, PIR, physical activity (PA), screen time (ST), and serum vitamin D. Subgroup analyses and interaction tests were performed for categorical covariates: sex, race and ethnicity, PIR, educational level of guardians, screen time and physical activity. In addition, a restricted cubic spline (RCS) curve was displayed to test the potential nonlinearity between BRI and myopia. Finally, to assess the robustness of association results, sensitivity analyses were performed using weighted ordinal logistic regression to assess whether the impact of BRI on different degrees of myopia remained consistent and using the waist-height ratio (WHtR) as exposure to explore the association with myopia.

All statistical analyses were performed using the statistical package R version 4.3.2 (http://www.R-project.org, The R Foundation). Statistical significance was established with a two-sided *P* value of <0.05.

## 3. Results

### 3.1 Characteristics of the participants

Among 3915 eligible adolescents with complete data, the weighted mean age (SE) was 14.37(0.04) years, and 2016 (50.86%) were women. The weighted proportions of adolescents with emmetropes, mild myopia, moderate myopia, and high myopia were 55.84%, 33.64%, 8.50%, and 2.02%, respectively. The characteristics of the participants are shown in Table 1 according to their BRI. We classified BRI into quartiles: Q1,< 2.083; Q2, 2.083 to 2.724; Q3, 2.725 to 4.004; Q4, ≥ 4.005. There were statistically significant differences in age, sex, race/ethnicity, PIR, educational level of guardians, WC, height, WHtR, physical activity and VD among different BRI groups (*P* < 0.05).

**Table 1.**
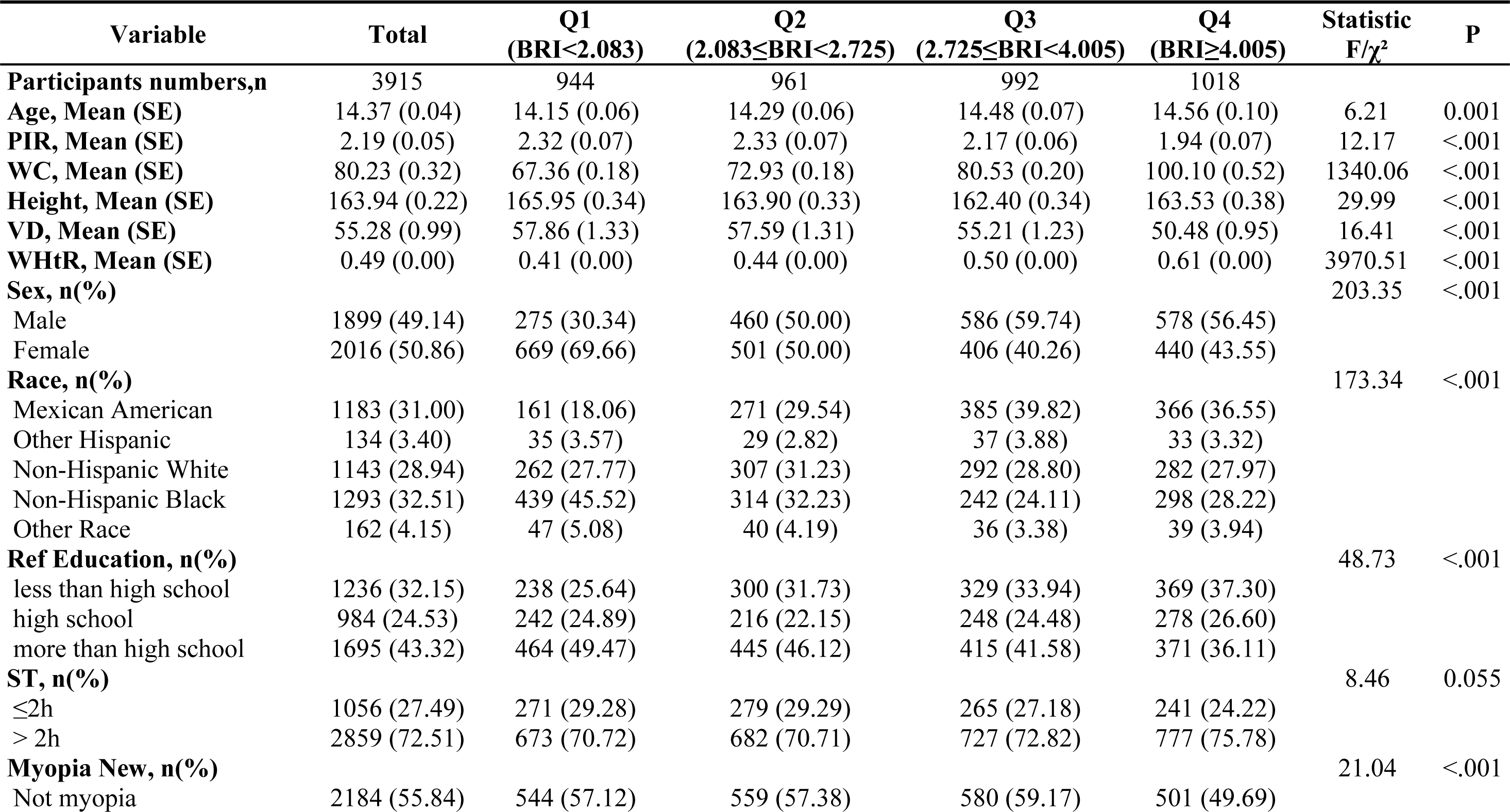
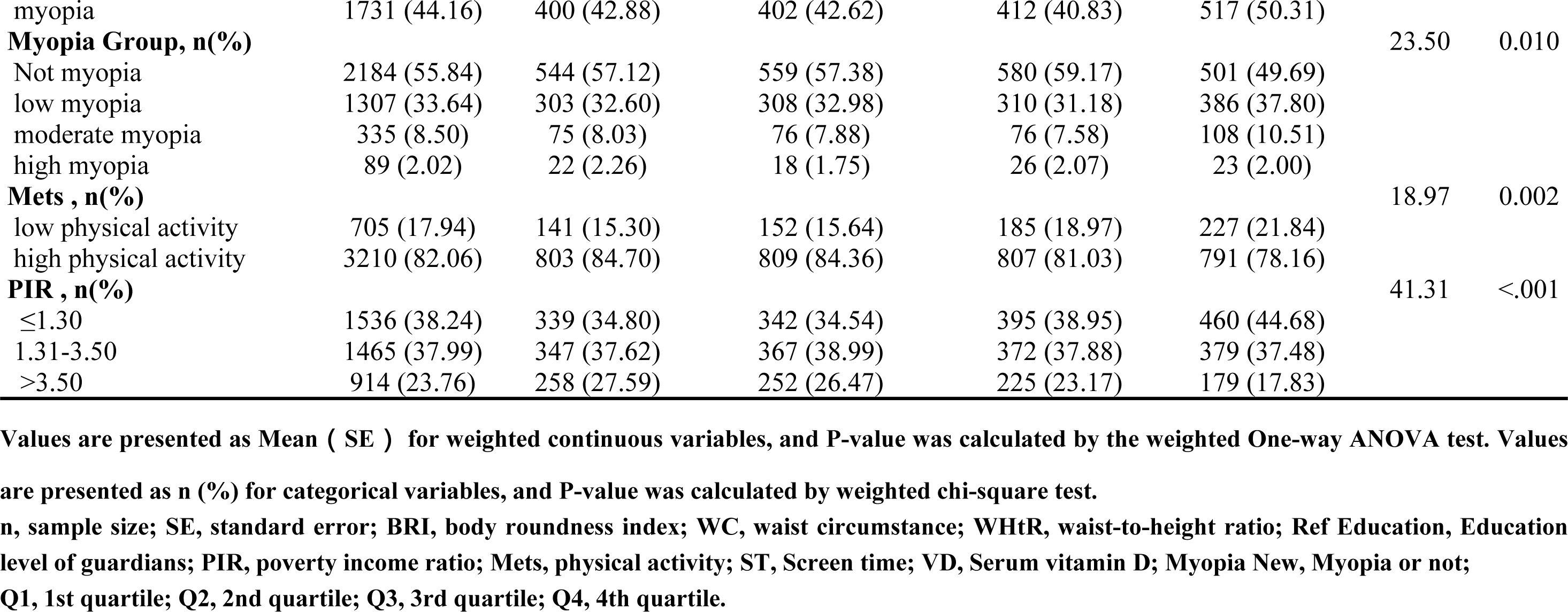
Baseline characteristics of the study participants grouped by quartiles of the BRI.

Adolescents who are older, have a larger waist circumference, a lower level of vitamin D, a relatively low PIR level and a higher WHtR are more likely to have a higher BRI. Adolescents with the highest BRI in Q4 are those whose guardians have a low educational level (less than high school) and a higher proportion of screen time of more than 2 hours, representing approximately 75.78%. The proportion of participants in Q4 with low physical activity was 21.84%, significantly higher than the other three groups.

### 3.2 Association between BRI and myopia

Weighted univariate and multivariate logistic regression analyses were used to demonstrate the relationship between BRI and myopia. Given that so far no BRI cut-off points were recommended, the BRI was categorized into quartiles (Q1, lowest, to Q4, highest) and Q1 (<2.083) was assigned as the reference group. The associations of BRI in categories with myopia before and after adjustment for covariates are provided in Table 2. We discovered a positive association between BRI and myopia. In the crude model, BRI was positively correlated with myopia (odds ratio [OR] = 1.057, 95%confidence interval [CI]: 1.016– 1.100, *P*<0.01). After adjustment for covariates, there were also significant relationships between BRI and myopia in model 2-3. In Model 3, for each one-unit increase in BRI, the incidence of myopia increased by 4.9% (OR = 1.049, 95%CI: 1.009-1.090, *P*=0.022). Specifically, after full adjustment for age, sex, race/ethnicity, PIR, Educational level of guardians, VD, Mets, ST, adolescents in Q4 were 26.1% more likely to develop myopia compared to adolescents in Q1 (OR=1.261, 95% CI: 1.046-1.521, *P*=0.022); findings were similar for adolescents in Model 2 (OR=1.255; 95% CI: 1.036-1.522), and Model 1 (OR=1.349, 95%CI:1.16-1.630). After adjusting for potential confounders, participants in the highest BRI quartile (Q4) showed a positive correlation with myopia compared to the lowest quartile(Q1) with P<0.05. In all models, the results indicated an overall linear trend between BRI and myopia. (All *P*-values for trend were significant).

**Table 2.** Weighted logistic regression analysis on the association between BRI and myopia.

|  | **Model 1** |  | **Model 2** |  | **Model 3** |  |
| --- | --- | --- | --- | --- | --- | --- |
|  | **OR (95% CI)** | **P Value** | **OR (95% CI)** | **P Value** | **OR (95% CI)** | **P Value** |
| **BRI continuous** | 1.057 (1.016 ~ 1.100) | 0.009 | 1.049 (1.008 ~ 1.090) | 0.023 | 1.049 (1.009 ~ 1.090) | 0.022 |
| **BRI-Q1** | Reference | - | Reference | - | Reference | - |
| **BRI-Q2** | 0.989 (0.834 ~ 1.172) | 0.900 | 0.941 (0.791 ~ 1.119) | 0.496 | 0.936 (0.790 ~ 1.109) | 0.452 |
| **BRI-Q3** | 0.919 (0.740 ~ 1.141) | 0.448 | 0.843 (0.672 ~ 1.059) | 0.151 | 0.837 (0.664 ~ 1.055) | 0.143 |
| **BRI-Q4** | 1.349 (1.116 ~ 1.630) | 0.004 | 1.255 (1.036 ~ 1.522) | 0.026 | 1.261 (1.046 ~ 1.521) | 0.022 |
| **P for trend** |  | 0.002 |  | 0.006 |  | 0.005 |

Data are presented as OR (95% CI). Model 1: no adjustment; Model 2: adjusted for age, sex, race/ethnicity; Model 3: adjusted for age, sex, race/ethnicity PIR, Ref Education, VD, Mets, ST.
OR, odds ratio; CI, confidence interval; BRI, body roundness index; Ref Education, Education level of guardian; PIR, poverty income ratio; Mets, PA; ST, Screen time; VD, Serum vitamin D; Q1, 1st quartile; Q2, 2nd quartile; Q3, 3rd quartile; Q4, 4th quartile.

### 3.3 Subgroup analyses

The results of the subgroup analyses and interaction tests are summarized in Fig 2. Subgroup analyses were conducted using sociodemographic factors after full adjustment. Positive associations between BRI and myopia were observed in Mexican American adolescents (OR=1.11, 95% CI: 1.05-1.19, *P*=0.001), women (OR=1.10, 95%CI: 1.05-1.15, *P*<0.001), those whose guardians’ educational level was less than high school (OR=1.09, 95%CI: 1.03-1.14, *P*=0.004), those with PIR of less than1.30 (OR=1.07, 95%CI: 1.01-1.14, *P*=0.037) and those with high physical activity (OR=1.08, 95%CI: 1.04-1.13, *P*<0.001). Interaction effects were observed in sex and physical activity. (*P* for interaction <0.05).

**Figure 2.**
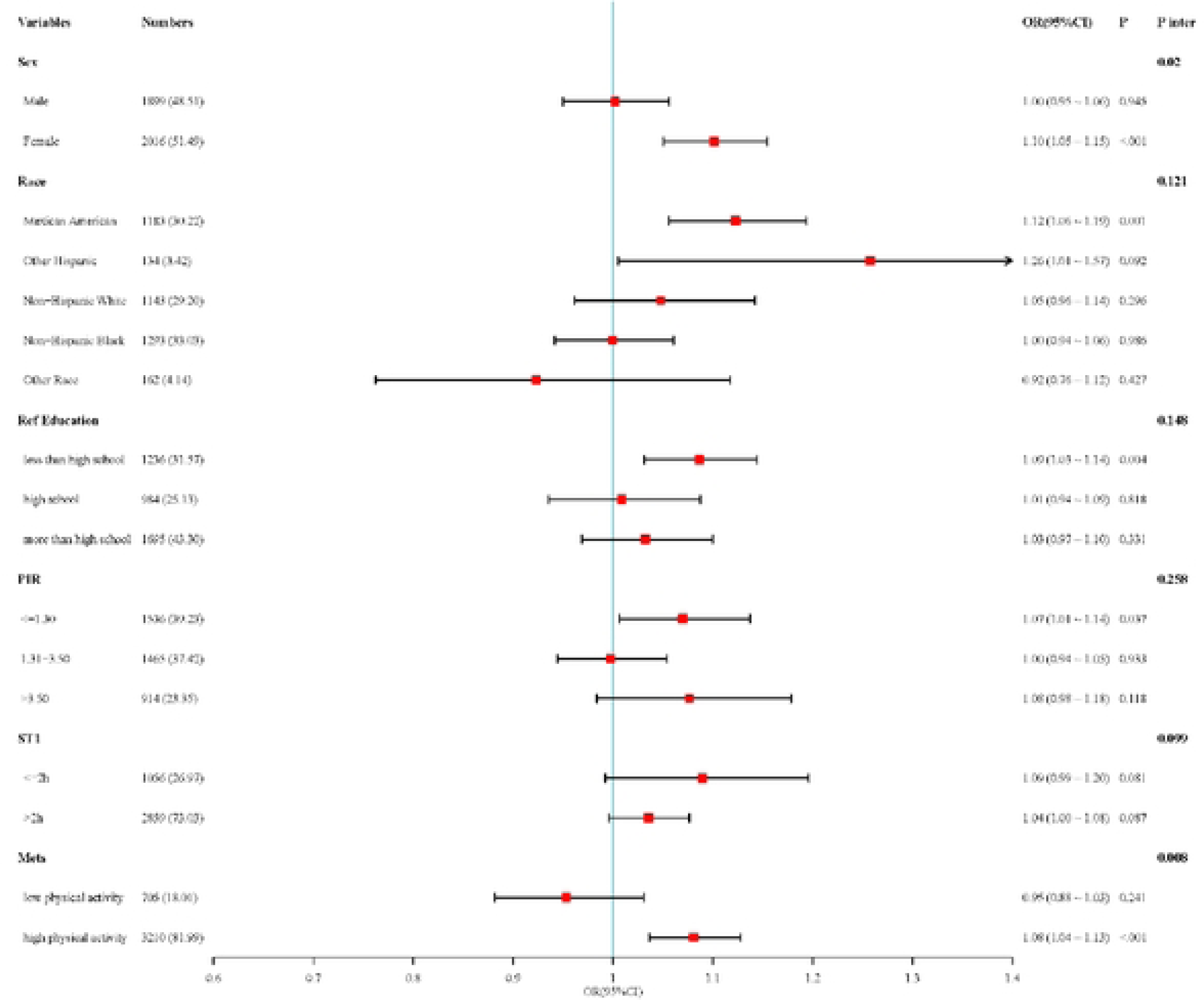
Subgroups analyses for the association between BRI and myopia. Model 3 was used in subgroup analysis, which was adjusted for adjusted for age, sex, race/ethnicity, PIR, Ref Education, VO, Mets, ST.OR, odds ratio; CI, confidence interval; BRI, body roundness index; Ref Education, Education level of guardian; PIR, poverty income ratio; Mets, PA; ST, Screen time; VO, Serum vitamin D; P inter, Interaction p-value.

### 3.4 Nonlinear correlation

In addition, the nonlinearity between BRI and myopia was resolved by restricted cubic splines (RCS). The results shown in Fig 3 revealed that there was an overall linear trend between BRI and myopia (all *P* values for trend <0.001) and nonlinear associations were shown in the three models (all *P* for nonlinear < 0.05), model 1 *P* for nonlinear=0.030, model 2 *P* for nonlinear=0.010 and model 3 *P* for nonlinear=0.007. Further analysis of the threshold effect showed that the inflection point for the BRI was 2.557.

**Figure 3.**
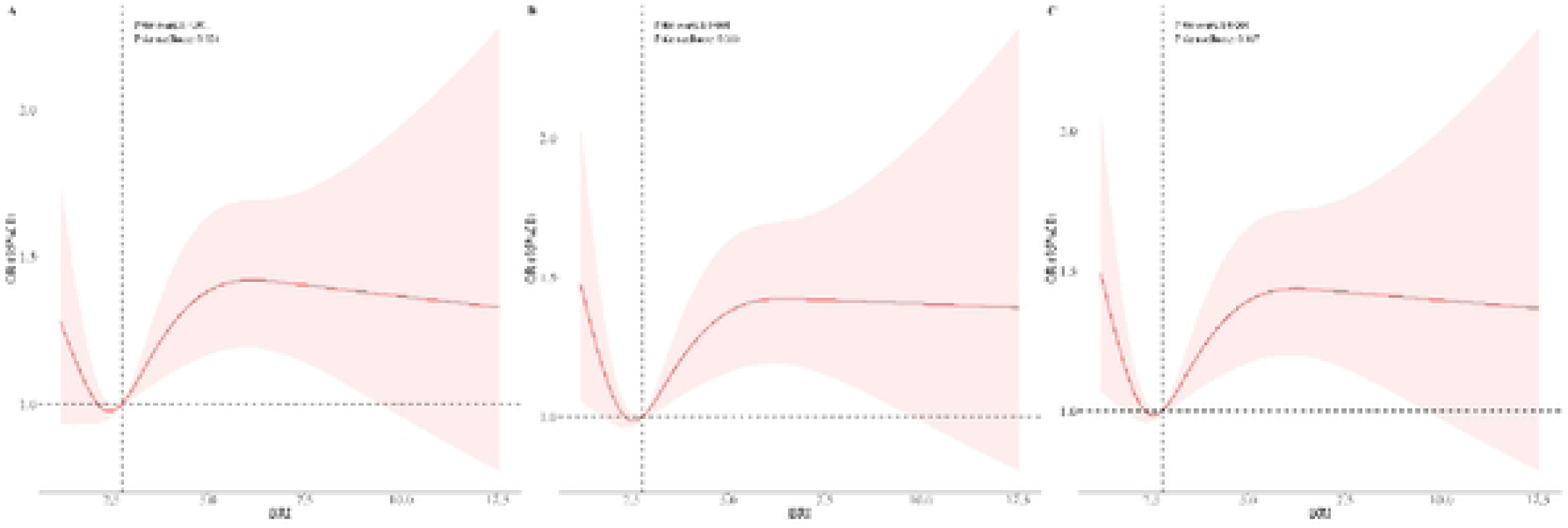
The RCS curve of the association between BRI and myopia among all study participants. (A) RCS regression was no adjustment; (B) RCS regression was adjusted for age, sex, race/ethnicity; (C) RCS regression was adjusted for age, sex, race/ethnicity, PIR, Ref Education, VD, Mets, ST. OR, odds ratio; CI, confidence interval; RCS, restricted cubic spline; BRI, body roundness index; Ref Education, Education level of guardian; PIR, poverty income ratio; Mets, PA; ST, Screen time; VD, Serum vitamin D.

### 3.5 Sensitivity Analyses

Sensitivity analyses were performed to test the stability and extrapolation of the association results. Myopia was classified as mild, moderate, and high myopia. Weighted ordinal logistic regressions were applied to explore the relationship of BRI and the quartiles of BRI with degrees of myopia. The positive correlation between BRI and myopia remained in different models: Model 1 OR=1.05, 95%CI:1.01-1.10, *P*=0.013; Model 2 OR=1.05, 95%CI:1.01-1.09, *P*=0.029 and Model 3 OR =1.05, 95%CI:1.01-1.09, *P*=0.020). Wald tests were used to analyze the BRI quartiles and adolescent myopia, and the results showed significant positive correlations (all *P*<0.01, Model 1 *P*=0.001, Model 2 *P*=0.002 and Model 3 *P*=0.001). After adjusting for potential confounders, participants in the highest quartile of BRI (Q4) had a positive correlation with myopia compared to those in the lowest quartile (Q1). The results are shown in S2 table.

The waist-height ratio (WHtR) is a widely used biomarker for central obesity. It is low cost, safe and widely available, and has high predictive utility in the clinical setting [31]. WHtR is determined by dividing WC by height (both measured in centimeters). S3 table shows the relationship between WHtR and myopia by weighted logistic regression. There was also a positive association between WHtR and myopia, which remained stable after adjusting for potential confounders, consistent with the results of BRI and myopia.

## 4. Discussion

This cross-sectional study derived data from NHANES to explore the correlation between BRI and myopia in American adolescents. In our study, 3915 participants were included. We found that adolescents who were older, had a larger waist circumference, a lower level of vitamin D, a relatively low PIR level and a higher waist-height ratio were more likely to have a higher BRI. By multivariate logistic analysis, we observed a positive association between BRI and myopia in adolescents (OR= 1.057, 95% CI: 1.016–1.100, *P*<0.01). After adjustment for several covariates, the association remained significant. Furthermore, the risk of developing myopia increased 26.1% in adolescents with BRI in Q4 (BRI ≥ 4.005) than Q1 (BRI<2.083) (OR=1.261, 95%CI: 1.046-1.521, *P*=0.022). RCS showed a nonlinear relationship between BRI and myopia (all *P* for nonlinear <0.05).

To our knowledge, this is the first study to evaluate the association of BRI with myopia. Obesity is an underlying risk for vision problems. There are many other indicators representing obesity, such as BMI, weight, WC, and WHtR, among which BMI is the most representative measure. The relationship between BMI and myopia has been extensively studied, but the results are inconsistent. A large cross-sectional study conducted in Korea reported that a decrease in BMI was associated with an increased prevalence of myopia [32]. However, this study was conducted among 19-year-old males, excluding females, and categorized BMI into four groups (<18.5, 18.5-22.99, 23.0-24.99, and >25.0), different from commonly used(<18.5, 18.5-24.9, 25-29.9, and>29.9). On the contrary, several studies have shown that BMI was positively associated with myopia[17–19]. Qu Yaohui, et al. reported a positive linear relationship between BMI and myopia (OR 1.01; 95% CI 1.01-1.02; *P*<0.05) [17]. Similarly, a Korean study of 938 children aged 5 to 18 years found that a higher BMI was significantly associated with high myopia [19]. Similarly, in the current study, we found that an increase in BRI was associated with an increased risk of myopia in adolescents. Subgroup analysis and interaction tests corroborated when stratified by gender, the association remained significant in girls, while the effect in boys did not show significance, which was consistent with Wang Jingjing, et al [33].

The association between BRI and myopia is affected by the underlying lifestyle and genetic factors. In our study, we observed that a higher BRI was correlated with an increased risk of myopia, especially in female adolescents, indicating that there may be some differences in the pathogenesis of myopia between sexes. The participants in our study are adolescents aged 12-19years. One possible explanation is the effect of estrogen and growth hormone. Estrogen has a certain impact on cornea thickness and regulation of MMP-2 expression in the human retinal pigment epithelium, which was positively correlated with myopic diopter [34–36]. Female myopia occurs earlier and the degree gradually increases more with body growth and development than male. In addition, estrogen plays an important role in the development of obesity by stimulating adipocytes and adipose tissue to accumulate and perform adipocyte functions, directly or indirectly [37, 38]. Another reason for the gender difference in the relationship between BRI and myopia may attribute to different lifestyle factors, including fewer outdoor time [39], more sedentary time, and high glycemic load carbohydrate diets of girls [40, 41]. One more explanation is socioeconomic aspects. The PIR as a ratio of self-reported or proxy family income to the federal poverty level based on family size reflects economic conditions. Better economic conditions may lead to greater use of close eye during their education and academic achievements [42]. Families with high economic levels appear to have a higher prevalence of myopia in children. The findings on the relationship between parental education level or myopia were consistent with previous reports. Myopia was also found more frequently in those whose mothers or parents had a low educational status[42, 43]. The low educational status of the parents usually represents low socioeconomic factors and makes them pay little attention to the occurrence of myopia in their children. We also observed that Mexican Americans had a higher risk of myopia occurrence, potentially attributed to genetic differences [44]. The aforementioned factors may explain the mechanisms of higher BRI that increase the risk of myopia in adolescents.

In conclusion, our study assessed that in adolescents an increase in BRI was associated with an increased risk of myopia, especially in women. Additionally, there was a nonlinear association between BRI and myopia. BRI exhibited an important threshold effect on myopia. This study aimed to increase public awareness of BRI values, a novel measure of obesity, and that maintaining a moderate BRI can help reduce the risk of myopia.

### Strengths

Our study has several strengths. First, the sample for this study came from NHANES, a large nationally representative sample considering sample weights. We adjusted for multiple confounding variables and performed subgroup analyses to ensure that our findings were widely applicable. Second, this is the initial study aimed at exploring the association between the new obesity index BRI and myopia. Third, we conducted interaction tests and subgroup analysis to further assess the relationship between BRI and myopia, and RCS was used to test the nonlinear relationship and identify reliable thresholds, offering new insights for the prevention and control of myopia.

### Limitations

However, our research still has some limitations. First, this cross-sectional study cannot prove a causal association between BRI and myopia. Second, the database lacks information on the axial length, which is another important standard to measure the progression of myopia. Moreover, the refractive power has not been measured after cycloplegia. Therefore, the results are not accurate enough. Third, although we considered several confounding variables, information about near work time, outdoor time, etc., was not included in NHANES. This may lead to bias in the results. More relevant future studies are expected to validate our findings.

## Data Availability

This database can be accessed on the official website of NHANES: www.cdc.gov/nchs/NHANES.

